# Registry for vascular cognitive impairment treatment with traditional Chinese medicine (REVIEW-TCM): Rationale and design of a prospective, observational study

**DOI:** 10.1101/2023.05.24.23290492

**Authors:** Xie Yao, Le Xie, Junlin Jiang, Ting Yao, Guo Mao, Rui Fang, Fuliang Kang, Shiliang Wang, Anchao Lin, Ying Gao, Jinwen Ge, Dahua Wu

## Abstract

**Background:** Vascular cognitive impairment (VCI) is one of the most common diseases among the elderly. However, few effective drugs have been approved for VCI. Traditional Chinese medicine (TCM) has been used in dementia for thousands of years. Currently, there is limited high-quality evidence for the efficacy of TCM, and the specific characteristics of its effects and the appropriate patient populations for TCM therapies remain unclear. Herein, we aim to explore the effectiveness and safety of TCM by conducting a longitudinal, patient-centered study.

**Methods:** REgistry for Vascular cognitive Impairment trEatment With Traditional Chinese Medicine (REVIEW-TCM) is a prospective, observational disease registry study. 1000 VCI patients at the Hunan Hospital of Integrated Traditional Chinese and Western Medicine will be recruited based on the following criteria: aged 18 years or older, Montreal Cognitive Assessment (MoCA) score < 26, and Hachinski Ischemic Score (HIS)≥7. There is no strict limit on the intervention, and different TCM formulas will be focused. Cognition, activity of daily living, quality of life, mental, psychology, *ZHENG* of TCM, and burden of caregiver will be evaluated at admission, and 6, 12, 18, and 24 months. Meanwhile, biological tests and neuroimaging examination will be applied to further explore the mechanism of TCM. Especially, a mixed-methods embedded design will be applied by adopting quantitative and qualitative studies to explore patients-reported outcomes of TCM. Finally, propensity score matching will be adopted to analyze the effectiveness of TCM.

**Discussion:** To the best of our knowledge, the REVIEW-TCM study is the first comprehensive, prospective, mixed-methods, registry-based study to evaluate TCM treatment in VCI, which will analyze the effectiveness and safety of TCM in the real world and explore population characteristics and subtypes of VCI suitable for TCM.

**Study registration:** This study was registered on www.chictr.org.cn (ChiCTR2200064756).

## Introduction

Vascular cognitive impairment (VCI) refers to the entire spectrum of vascular brain pathologies that contribute to any degree of cognitive impairment [1]. VCI can be classified as mild or major, with the latter also being known as vascular dementia (VaD). Over the past few years, vascular mild cognitive impairment has become increasingly common, with a reported prevalence of around 42% in people over 65 years old [2]. In China, there are currently an estimated 2.49 million patients with vascular dementia [3]. It is well-established that patients with VCI can experience a range of cognitive and quality-of-life impairments. The commonly impaired cognitive domains in VCI patients involve executive function, attention, and processing speed [4], which can hinder processes for initiation, planning, decision-making, hypothesis generation, cognitive flexibility, and judgment. In addition, other clinical features such as gait disturbance, urinary incontinence, and emotional disorders can make work and daily life more arduous [5].

Due to the gradual decline in their cognitive abilities, individuals with VCI may become increasingly dependent on others for their activities of daily living, which can lead to a decline in their overall well-being. Experiencing anxiety and depression is very common in people with cognitive impairment [6], and emotional disorders can also affect the caregiver. Indeed, the caregiver burden will remarkably increase under caring for major cognitive impairment [7]. Accordingly, during the treatment of VCI, behavioral and psychological symptoms should be addressed, providing support for patients and caregivers and maximizing independence [8].

The current medical approach for VCI treatment often exhibits limited efficacy, as cholinesterase inhibitors and memantine provide minimal benefits [9]. Traditional Chinese medicine (TCM) has been used in dementia for thousands of years. There is an increasing consensus that Chinese medicine is beneficial for VCI [10-13]. Many studies have shown positive effects in improving cognitive impairment, psychological symptoms and quality of life [14]. However, there is limited high-quality evidence for TCM, and it remains uncertain which patient populations and therapeutic approaches are most appropriate and effective. Meanwhile, the course of treatment and effective combination therapies warrant further investigation [15].

Indeed, a prospective observational study with regular and long-term follow-ups can address the above questions. Registry for vascular cognitive impairment treatment with traditional Chinese medicine (REVIEW-TCM) aims to investigate the characteristics and effectiveness of TCM for VCI in the Chinese population. The key factors affecting the clinical effectiveness and suitable population characteristics for TCM will be explored. In addition, the neuroimaging scan images and blood samples will be collected for further mechanism research. Finally, the perspective of patients and caregivers will be analyzed to explore patient-reported outcomes of TCM.

## Methods and design

### Study design

The REVIEW-TCM study (registered with www.chictr.org.cn, ChiCTR2200064756) is a prospective observational disease registry that utilizes a mixed-methods approach, incorporating both quantitative and qualitative study designs. Figure 1 and Figure 2 provide an overview of the study. The research protocol has been approved by Hunan Hospital of Integrated Traditional Chinese and Western Medicine, Changsha, China, on 20 July, 2022. (No. [2022] 48).

**Figure 1.**
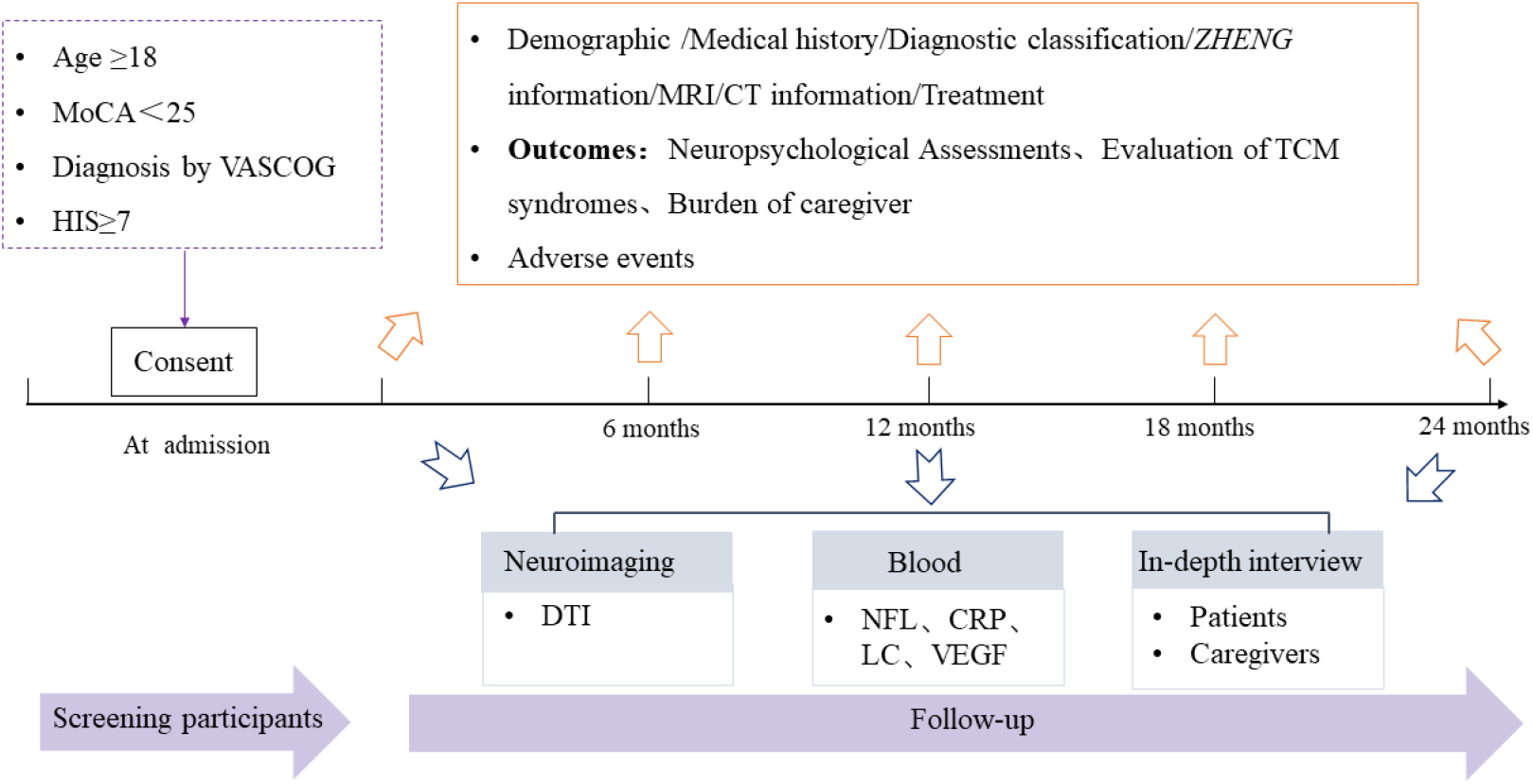
The overview of study flowchart of REVIEW-TCM CT, Computerized tomography; CRP, C-reactive protein; DTI, Diffusion tensor imaging; HIS, Hachinski Ischemic Score; LC, Leukocyte count; NFL, Neurofilament light chain; MoCA, Montreal Cognitive Assessment; MRI, Magnetic resonance imaging; REVIEW-TCM, Registry for vascular cognitive Impairment treatment with traditional Chinese medicine; TCM, Traditional Chinese medicine; VASCOG, The special symposium of the International Society for Vascular Behavioral and Cognitive Disorders; VEGF, Vascular endothelial growth factor.

**Figure 2.**
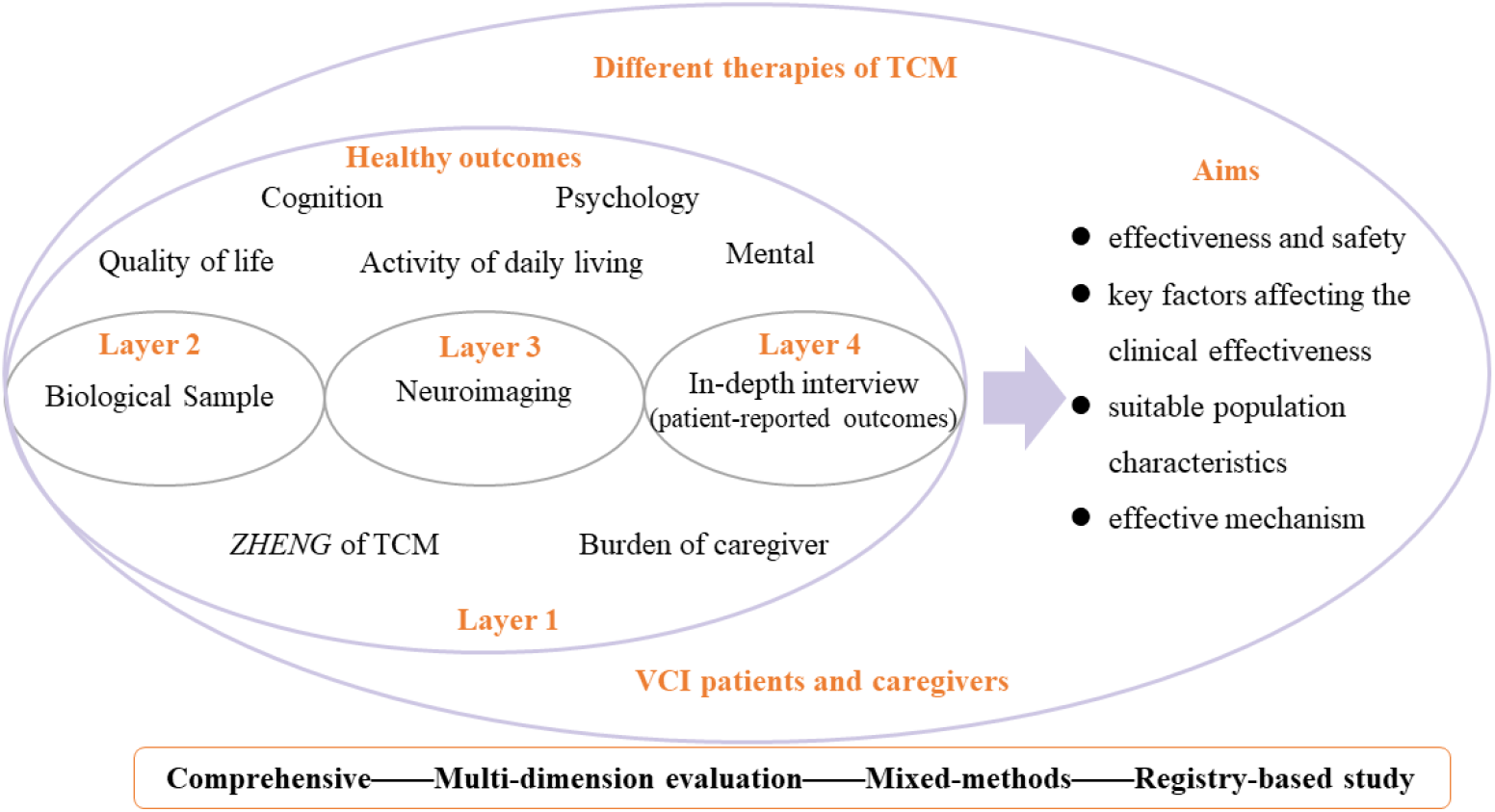
Study design of registry for REVIEW-TCM

### Patient population

Patients aged 18 years or older were consecutively recruited from Hunan Hospital of Integrated Traditional Chinese and Western Medicine, from October 30, 2022 to December 31, 2027. The special symposium of the International Society for Vascular Behavioral and Cognitive Disorders (VASCOG) [16] will be adopted to diagnose VCI, the Montreal Cognitive Assessment (MoCA) score should be lower than 26, and the Hachinski Ischemic Score (HIS) should be greater than 6. The exclusion criteria are as follows: (1) patients diagnosed with Alzheimer’s disease, frontotemporal dementia, Lewy body dementia, Parkinson’s disease, multiple sclerosis, brain trauma and other neurological diseases leading to cognitive impairment; (2) patients with altered levels of consciousness, unstable vital signs or serious cardiopulmonary diseases; (3) patients unable to complete the study due to psychological or emotional disorders, severe audio-visual disorders. All participants or proxies should give informed consent.

### Specific treatment

There is no strict limitation placed on the intervention. We will focus on TCM treatment and pay attention to the detailed information, including herbs in prescriptions, formulation (Chinese patent medicine and TCM decoctions), duration (start and stop times), and therapy compliance. The rest of the treatment protocol are according to guidelines, including donepezil, galantamine, rivastigmine, memantine, butylphthalide, citicoline, cognitive rehabilitation training, etc [4].

### Neuropsychological Assessments

The cognitive scales selected are based on the recommendations from the vascular impairment of cognition classification consensus study (VICCCS) [17]. MoCA will be used to assess general cognitive function. Trail Making Test A (TMT-A) and Test B (TMT-B) will be applied to measure patients’ processing speed and executive function. The verbal learning, recognition memory, and recall will be evaluated by Hopkins Verbal Learning Test (HVLT). Language proficiency will be assessed using Boston Naming Test, 2nd Edition (BNT-2). Visuospatial abilities will be evaluated with the Clock Drawing Test (CDT).

Neuropsychiatric symptoms will be assessed using Neuropsychiatric Inventory (NPI). Anxiety and depression will be measured by Hospital Anxiety and Depression Scale (HADS). Disability will be assessed with Instrumental Activities of Daily Living (IADL). Health-related quality of life will be assessed by the European Quality of Life-5 Dimensions-5 Levels (EQ-5D-5L). The evaluators have accomplished standardized training and kept good internal consistency.

### Evaluation of TCM syndromes

The scale for the differentiation of syndromes of vascular dementia (SDSVD) will be applied to diagnose *ZHENG* and evaluate TCM’s efficacy, including seven *ZHENG* of TCM. The maximum score of each *ZHENG* in SDSVD is 30. If one TCM *ZHENG*’s score is at least 7, the diagnosis is established. We will choose the change scores after treatment to verify the clinical efficacy of TCM.

### Burden of caregiver

It is widely acknowledged that the caregivers who take care of VCI patients are significantly more stressed, which leads to a decline in the caregiver’s well-being. The caregiver-oriented outcome will be assessed using the Zarit Burden Interview (ZBI), with a total score of 88, involving 22 items. The role burden and personal burden will be assessed in ZBI.

### Biological Sample Tests

Blood samples will be collected from participants who give informed consent. However, participants diagnosed with infections, hematological disease, malignant tumors, and other neurological diseases will be excluded. The serum and plasma will be collected from ten milliliters of blood within 30min centrifuged at 3,000 r/min for 20 min. Then, 400ul serum or plasma in each polypropylene tube will be stored at -80°C. The neurofilament light chain (NFL), C-reactive protein (CRP), leukocyte count (LC), and vascular endothelial growth factor (VEGF) will be examined at regular intervals during the study. The blood samples will be acquired from fasting patients at each visit. (Supplementary Figure S1)

### Imaging Examination and Analysis

Magnetic resonance imaging (MRI) will be performed on consenting participants who are right-handed and have a HADS score < 8 and a modified Rankin scale (mRS) score ≤ 2. All participants should have no contraindications for MRI and without other neurological diseases or psychiatric disorders. MRI acquisitions will be carried on 3.0 T GE Signa Scanner. The standardized neuroimaging protocol includes three-dimensional T1-weighted imaging (T1WI), T2-weighted imaging (T2WI), magnetic resonance angiography (MRA), and diffusion tensor imaging (DTI) sequence. The infarct or hemorrhage lesions, white matter hyperintensity (WMH), vascular stenosis, hippocampal atrophy, mean diffusivity (MD), and fractional anisotropy (FA) will be recorded. The neuroimaging analysis will be conducted by two experienced radiologists independently. (Supplementary Figure S2)

### Patient-reported outcomes

To explore the potential therapeutic effects of TCM, we plan to interview participants (VCI patients and their caregivers) undergoing TCM treatment for at least 3 months. The score of MoCA in VCI patients should be between 10-26, and cognitive impairment is their main symptom. All participants will be assessed by a 30 min semi-structured questionnaire, including disturbing symptoms, emotional needs, experience of TCM, etc. The grounded analysis will be used to collect items on treatment effects and generate conceptual graphs, which will serve as the basis for developing patient-centered outcomes. Finally, the Delphi survey and verification research will be conducted to obtain patient-reported outcomes [18] (Supplementary Figure S3).

### Data collection and monitoring

All data will be collected on an electronic data capture (EDC) system (http://www.1-dao.net) with unique ID during the study period. The main data elements include demographics, diagnosis, cognition, examination, imaging, treatments, outcomes, safety information, and other information (Table 1). Informed consent requires scanning and uploading. The EDC system can remind researchers of the time to follow up and automatically check for completeness and logical error in the uploaded data.

**Table 1.**
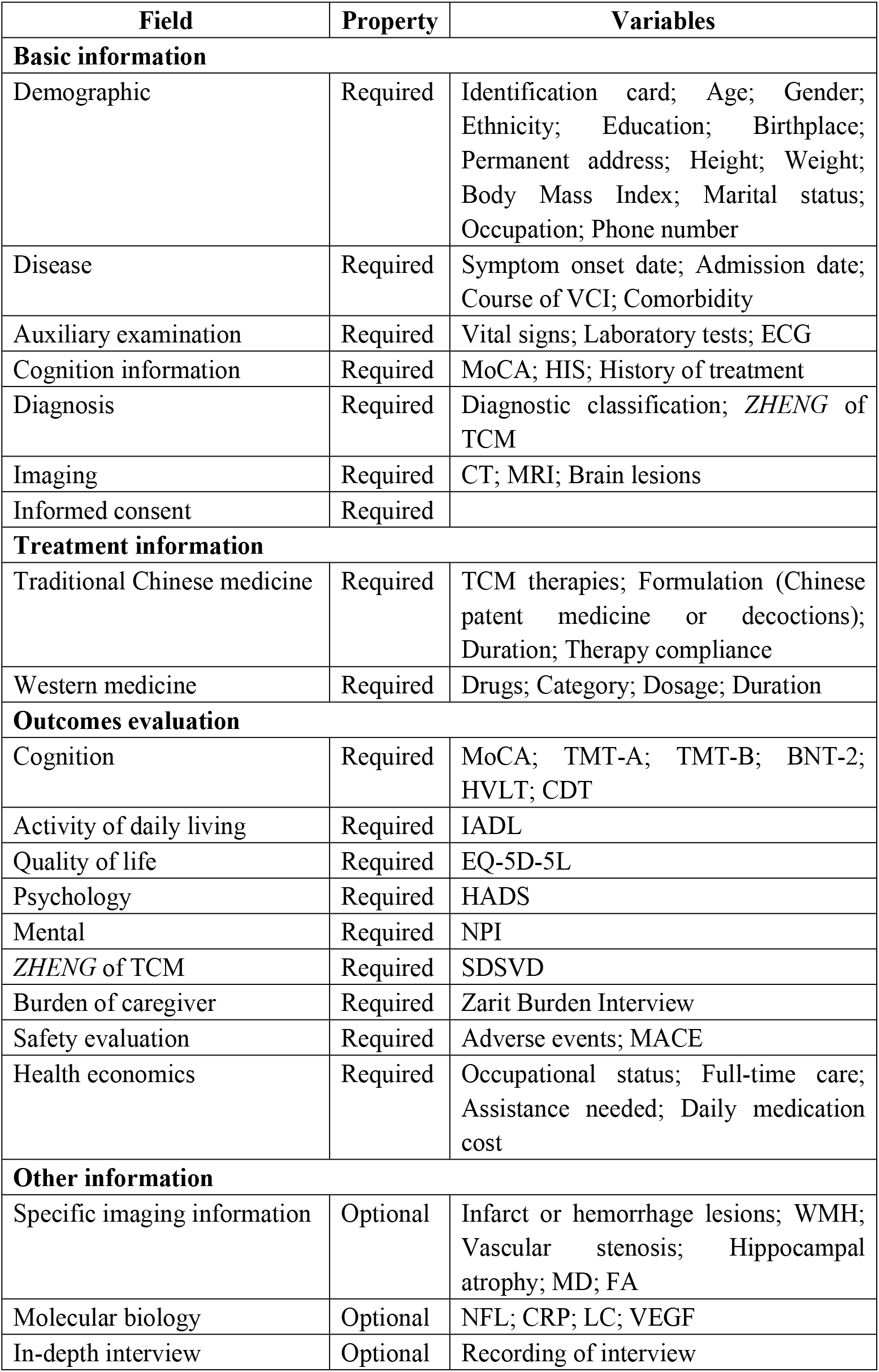
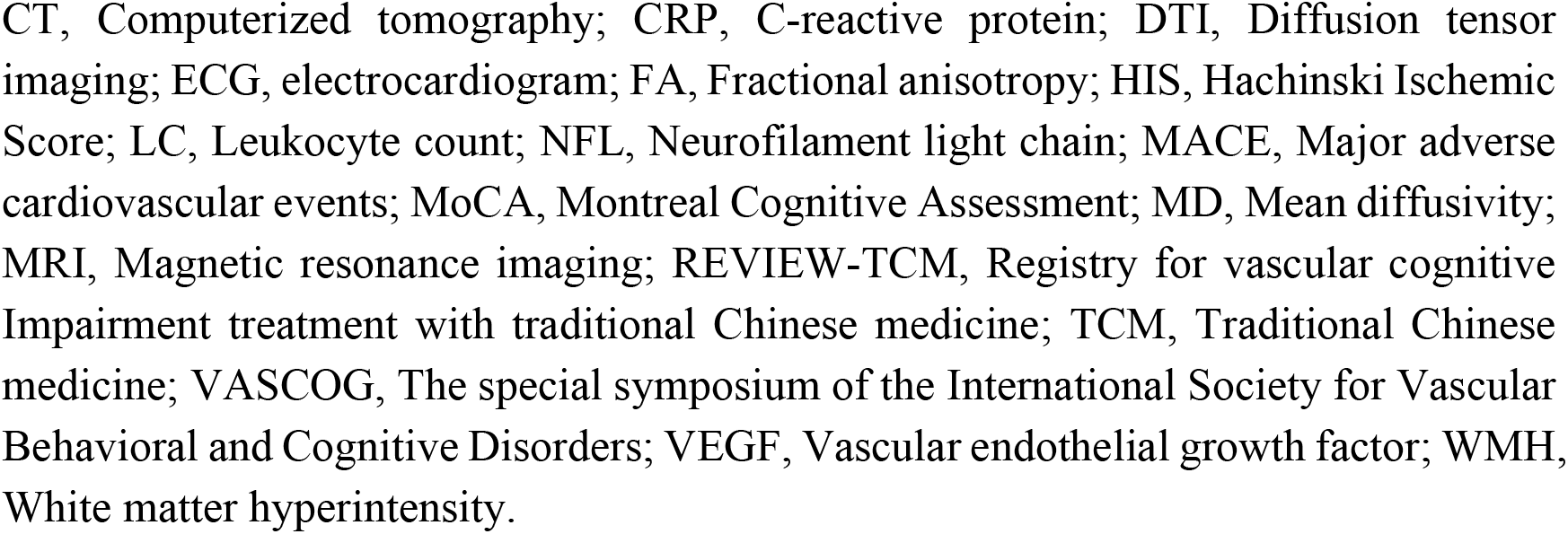
Main variables registered in REVIEW-TCM study

| Field | Property | Variables |
| --- | --- | --- |
| **Basic information** |  |  |
| Demographic | Required | Identification card; Age; Gender; Ethnicity; Education; Birthplace; Permanent address; Height; Weight; Body Mass Index; Marital status; Occupation; Phone number |
| Disease | Required | Symptom onset date; Admission date; Course of VCI; Comorbidity |
| Auxiliary examination | Required | Vital signs; Laboratory tests; ECG |
| Cognition information | Required | MoCA; HIS; History of treatment |
| Diagnosis | Required | Diagnostic classification; *ZHENG* of TCM |
| Imaging | Required | CT; MRI; Brain lesions |
| Informed consent | Required |  |
| **Treatment information** |  |  |
| Traditional Chinese medicine | Required | TCM therapies; Formulation (Chinese patent medicine or decoctions); Duration; Therapy compliance |
| Western medicine | Required | Drugs; Category; Dosage; Duration |
| **Outcomes evaluation** |  |  |
| Cognition | Required | MoCA; TMT-A; TMT-B; BNT-2; HVLT; CDT |
| Activity of daily living | Required | IADL |
| Quality of life | Required | EQ-5D-5L |
| Psychology | Required | HADS |
| Mental | Required | NPI |
| *ZHENG* of TCM | Required | SDSVD |
| Burden of caregiver | Required | Zarit Burden Interview |
| Safety evaluation | Required | Adverse events; MACE |
| Health economics | Required | Occupational status; Full-time care; Assistance needed; Daily medication cost |
| **Other information** |  |  |
| Specific imaging information | Optional | Infarct or hemorrhage lesions; WMH; Vascular stenosis; Hippocampal atrophy; MD; FA |
| Molecular biology | Optional | NFL; CRP; LC; VEGF |
| In-depth interview | Optional | Recording of interview |

Any data correction and an electronic audit trail with an electronic signature and data will be recorded on the system. To ensure the security of participant data, the EDC system has implemented a strict authentication process.

### Follow-up procedures

Participants will be scheduled for face-face visit every three months over two years. Researchers will provide specialized health services to ensure study compliance and keep in touch with participants and their caregivers by phone monthly. The treatment information, physical examination, four examinations of TCM, outcomes evaluation, safety assessment, and major adverse cardiovascular events (MACE) will be collected at each visit (at admission and at 6, 12, 18, and 24 months). Blood sample collection and MRI examination will be performed annually.

### Study outcomes

To clarify the characteristics of the clinical efficacy of TCM therapies, the outcome will be evaluated from multiple dimensions, including cognition, the activity of daily living, quality of life, mental, psychology, *ZHENG* of TCM, the burden of caregiver, and adverse events. The primary outcomes are the change scores of MoCA from baseline to 12 months and 24 months. The secondary outcomes are the variation of MoCA, TMT-A, TMT-B, HVLT, BNT-2, CDT, NPI, HADS, IADL, EQ-5D-5L, SDSVD, ZBI from baseline to 12 and 24 months. The secondary outcomes will also include the incidence of adverse events and MACE during the study. The exploratory outcomes will be the change in NFL, CRP, LC, VEGF, MD, and FA.

### Quality control and management

Prior to study initiation, a committee will design standard operating procedures regarding consecutive patient screening and recruitment, obtaining informed consent, and data collection. Research personnel, statistical analysts, and data administrator should formulate the data verification plan together. Data administrators will regularly review the uploaded data according to the verification plan, such as missing data, logical errors, etc. Any questions about the uploaded data will be directed to the investigator for clarification. The data administrators will be required to produce a monthly data audit report.

### Study sample size

Currently, there is no valid sample size calculation method for registry studies. Multivariate analysis will be performed for the study outcomes. Based on the literature and expert opinions, the preliminary research variables will be no more than 40.

According to the rule of thumb [19-20], the principle of sample size estimation should be 20 times that of variables. Assuming a 20% dropout rate after the study is completed, a total sample size of 1000 participants will be needed.

### Statistical analysis

SPSS version 26 (IBM Corp., Armonk, NY, USA) will be applied for statistical analysis. The baseline characteristics of all participants will be described as follows: the mean, median, standard median, or interquartile ranges will be used to summarize continuous data, while counts or percentages for categorical data. Between-group comparisons will be conducted using ANOVA or t-test, or non-parametric tests.

The propensity score matching will be adopted to analyze the effectiveness of TCM. Baseline variables will be balanced and matched, such as age, sex, education, hypertension, diabetes, smoking, stroke, cognition, brain lesion, etc. The comparative analysis will be conducted after the baseline unevenness adjustment. Propensity score matching will be used for comparisons, such as different therapies (TCM vs. western medicine vs. combined treatment), formulations (Chinese patent medicine vs. decoctions vs. formula granule), etc.

Hierarchical models based on the cognitive impairment grading, brain lesion size, age group, gender, TCM syndromes, classification of diseases, course of treatment will be taken into account to explore important factors affecting the effectiveness of TCM for VCI. Conditional logistic regression models will be adjusted by different variables to identify predictors of long-term effectiveness presented by MoCA and other outcomes at 12- and 24-month follow-ups.

While quality control can improve the integrity of research data, missing data is often unavoidable in observational studies. However, limiting missing data to less than 20% is important. By convention, nonresponse follow-up will be classified as missing not at random (MNAR) [21]. Therefore, multiple imputations will be used to handle missing data [22].

## Discussion

Given the diagnostic and treatment challenges of dementia, an increasing number of observational studies have been conducted around the world, including the English Longitudinal Study of Aging (ELSA) [23], the German Aging Survey (DEAS) [24], and China Registry Study on Cognitive Impairment in the Elderly (HOPE) [25], with few studies on VCI. A registry study is an organized system that collects medical data in an observational format to evaluate clinical outcomes for specific diseases, health conditions, or exposed populations [26]. To our knowledge, only one TCM registry study [27] has focused on VCI, which just pay attention to one Chinese patent drug for vascular dementia. Generally speaking, these data lack comprehensive TCM information and multi-dimensional outcome assessment. The REVIEW-TCM study will explore the efficacy characteristics, suitable patient populations, and patient-centered outcomes of TCM therapies. It is worth noting that specific studies with hypotheses, such as comparative effectiveness research, pragmatic randomized clinical trials, or nested case-control studies, can be included in this registry study. This approach can lead to the generation of secondary hypotheses, which can be further tested in clinical trials [28].

TCM treatment is mainly based on syndrome differentiation. *ZHENG* of TCM can categorize different characteristics of VCI patients. According to *ZHENG*, different Chinese patent medicine and TCM decoctions can be prescribed by doctors. For example, Qi Fu Yin can tonify qi and replenish blood to boost qi and blood circulation, which includes bioactive compounds or phytochemicals to produce antioxidant, anti-inflammatory, and anti-thrombotic effects [29]. Therefore, Chinese herbs based on different *ZHENG* can influence different pathological mechanisms and VCI populations. Moreover, there is high-quality evidence supporting the use of Sailuotong (SLT) for tonifying qi and activating blood [11,13]. SLT is reportedly effective against VCI populations with mild to moderate severity dementia and evidence of ischemic lesions, improving functioning in multiple domains, such as memory, orientation, language and executive function [13]. Additionally, we sought to determine the specific characteristics of TCM treatments that are most effective in treating VCI, as well as the patient populations that are most likely to benefit from them.

Holism is a key principle of TCM which involves pursuing unity and integrity of function in humans and keeping harmony with the social environment [30]. Besides cognitive impairment, gait disturbance, urinary incontinence, emotional disorder, low quality of life, and social role can impact VCI patients and caregivers. The REVIEW-TCM study will pay attention to cognitive functions, the changes in ADL, global clinical improvement, quality of life, neuropsychiatric symptoms, and the burden of the caregiver. However, some VCI patient-centered outcomes may not be found for TCM treatment. Patient-reported outcomes will be explored in mixed methods research, which integrates qualitative and quantitative methods for scientific inquiry and evaluation to supplement outcomes [31]. Mixed methods research has been widely used in real-world studies to comprehensively answer research questions, providing abundant data for the individualized therapeutic evaluation of TCM in different aspects, which may reflect holism and humanities characteristics of TCM at the individual level. [32-33]

An increasing body of evidence suggests that white matter (WM) plays an important role in cognitive decline and dementia [34]. DTI is a kind of MRI technology to detect microscopic structural changes in WM and evaluate the structural integrity of WM fibers. It has become a potential candidate imaging marker to evaluate cognitive impairment [35]. Accordingly, we will pay more attention to DTI in neuroimaging examination to explore the effects of TCM on our brain structure. NFL is an intermediate filament protein that is a component of the cytoskeleton of neurons and is abundantly expressed in axons [36]. It is also a possible prognostic biomarker for VCI and may serve as a pharmacodynamic response biomarker for therapeutic target engagement [37]. Hypertension, diabetes, or other vascular risk factors can lead to cerebrovascular injury. In contrast, angiogenesis and systemic inflammation biomarkers, like VFGF, LC or CRP, may play a pathogenic role in driving cognitive decline [38-39]. These biomarkers will improve our understanding of the pharmacological mechanisms of TCM and represent potential therapeutic response indicators.

It has long been established that TCM treatment can boost memory and cognitive functions and manage behavioral and psychological symptoms associated with VCI. We conducted a longitudinal, patient-centered study to comprehensively collect epidemiological and clinical data for TCM research. To the best of our knowledge, the REVIEW-TCM study is the first comprehensive, prospective, mixed-methods, registry-based study to evaluate TCM treatment in VCI, which will analyze differences among different TCM therapies and the effectiveness and safety of TCM in the real world and explore population characteristics and subtypes of VCI suitable for TCM.

## Data Availability

No datasets were generated or analysed during the current study. All relevant data from this study will be made available upon study completion.

https://www.chictr.org.cn

## Acknowledgments

We gratefully acknowledge the contributions of the researchers and all participants in this study. We also thank Professor Ying Gao, Jinwen Ge, and DahuaWu for providing support on EDC in REVIEW-TCM.

## Authors’ contributions

The idea of this research was proposed by YX. This protocol drafting was undertaken by YX, LX, RF. DHW and JWG revised the manuscript for intellectual content. YG will provide EDC system. Data collection and security will be performed by YX, JLJ, GM, TY, FLK, SLW, ACL. All authors approved the final version of the manuscript.

## Availability of data and materials

Not applicable

## Declarations

### Ethics approval and consent to participate

This study protocol involving human participants has been reviewed and approved by Hunan Hospital of Integrated Traditional Chinese and Western Medicine, Changsha, China (No. [2022] 48). The study will be conducted in accordance with the principles of Good Clinical Practice and the Declaration of Helsinki. The study has also been registered at the Chinese Clinical Trial Registry (ChiCTR2200064756). All participants must give written informed consent which should be acquired after a sufficient and detailed explanation of the research situation (including confidentiality, risk and benefits, the purpose of the study, etc.).

### Consent for publication

Not applicable.

## References

1 van der Flier WM, Skoog I, Schneider JA, Pantoni L, Mok V, Chen C, et al. Vascular cognitive impairment. Nat Rev Dis Primers. 2018;4:18003. http://doi.org/10.1038/nrdp.2018.3

2 Jia J, Zhou A, Wei C, Jia X, Wang F, Li F, et al. The prevalence of mild cognitive impairment and its etiological subtypes in elderly Chinese. Alzheimers Dement. 2014;10(4):439–47. http://doi.org/10.1016/j.jalz.2013.09.008

3 Jia L, Quan M, Fu Y, Zhao T, Li Y, Wei C, et al. Dementia in China: epidemiology, clinical management, and research advances. Lancet Neurol. 2020;19(1):81–92. http://doi.org/10.1016/S1474-4422(19)30290-X

4 Cognitive Impairment Committee of Neurology Branch of Chinese Medical Doctor Association, Chinese Guidelines for the diagnosis and treatment of vascular cognitive Impairment 2019. National Medical Journal of China 2019; 99(35): 2737–2744. http://doi.org/10.3760/cma.j.issn.0376-2491.2019.35.005

5 Litak J, Mazurek M, Kulesza B, Szmygin P, Litak J, Kamieniak P, et al. Cerebral Small Vessel Disease. Int J Mol Sci. 2020;21(24). http://doi.org/10.3390/ijms21249729

6 Orgeta V, Qazi A, Spector AE, Orrell M. Psychological treatments for depression and anxiety in dementia and mild cognitive impairment. Cochrane Database Syst Rev. 2014;2014(1):D9125. http://doi.org/10.1002/14651858.CD009125.pub2

7 Dauphinot V, Potashman M, Levitchi-Benea M, Su R, Rubino I, Krolak-Salmon P. Economic and caregiver impact of Alzheimer’s disease across the disease spectrum: a cohort study. Alzheimers Res Ther. 2022;14(1):34. http://doi.org/10.1186/s13195-022-00969-x

8 Dichgans M, Leys D. Vascular Cognitive Impairment. Circ Res. 2017;120(3):573–91. http://doi.org/10.1161/CIRCRESAHA.116.308426

9 Iadecola C, Duering M, Hachinski V, Joutel A, Pendlebury ST, Schneider JA, et al. Vascular Cognitive Impairment and Dementia: JACC Scientific Expert Panel. J Am Coll Cardiol. 2019;73(25):3326–44. http://doi.org/10.1016/j.jacc.2019.04.034

10 Xu QQ, Shan CS, Wang Y, Shi YH, Zhang QH, Zheng GQ. Chinese Herbal Medicine for Vascular Dementia: A Systematic Review and Meta-Analysis of High-Quality Randomized Controlled Trials. J Alzheimers Dis. 2018;62(1):429–56. http://doi.org/10.3233/JAD-170856

11 Chang D, Liu J, Bilinski K, Xu L, Steiner GZ, Seto SW, et al. Herbal Medicine for the Treatment of Vascular Dementia: An Overview of Scientific Evidence. Evid Based Complement Alternat Med. 2016: 7293626. http://doi.org/10.1155/2016/7293626

12 Chan ES, Bautista DT, Zhu Y, You Y, Long JT, Li W, et al. Traditional Chinese herbal medicine for vascular dementia. Cochrane Database Syst Rev. 2018;12(12):D10284. http://doi.org/10.1002/14651858.CD010284.pub2

13 Jia J, Wei C, Chen S, Li F, Tang Y, Qin W, et al. Efficacy and safety of the compound Chinese medicine SaiLuoTong in vascular dementia: A randomized clinical trial. Alzheimers Dement (N Y). 2018;4:108–17. http://doi.org/10.1016/j.trci.2018.02.004

14 Zeng L, Zou Y, Kong L, Wang N, Wang Q, Wang L, et al. Can Chinese Herbal Medicine Adjunctive Therapy Improve Outcomes of Senile Vascular Dementia? Systematic Review with Meta-analysis of Clinical Trials. Phytother Res. 2015;29(12):1843–57. http://doi.org/10.1002/ptr.5481

15 Bai X, Zhang M. Traditional Chinese Medicine Intervenes in Vascular Dementia: Traditional Medicine Brings New Expectations. Front Pharmacol. 2021;12:689625. http://doi.org/10.3389/fphar.2021.689625

16 Sachdev P, Kalaria R, O’Brien J, Skoog I, Alladi S, Black SE, et al. Diagnostic criteria for vascular cognitive disorders: a VASCOG statement. Alzheimer Dis Assoc Disord. 2014;28(3):206–18. http://doi.org/10.1097/WAD.0000000000000034

17 Skrobot OA, O’Brien J, Black S, Chen C, DeCarli C, Erkinjuntti T, et al. The Vascular Impairment of Cognition Classification Consensus Study. Alzheimers Dement. 2017;13(6):624–33. http://doi.org/10.1016/j.jalz.2016.10.007

18 Xue-fei W, Qiang L, Hai-zhen Z, Ying G. Development process of patient-reported outcome draft scale of stroke. China Journal of Traditional Chinese Medicine and Pharmacy 2012; 27(02): 292–295.

19 van der Ploeg T, Austin PC, Steyerberg EW. Modern modelling techniques are data hungry: a simulation study for predicting dichotomous endpoints. Bmc Med Res Methodol. 2014;14:137. http://doi.org/10.1186/1471-2288-14-137

20 Riley RD, Ensor J, Snell K, Harrell FJ, Martin GP, Reitsma JB, et al. Calculating the sample size required for developing a clinical prediction model. BMJ. 2020;368:m441. http://doi.org/10.1136/bmj.m441

21 McPherson S, Barbosa-Leiker C, Mamey MR, McDonell M, Enders CK, Roll J. A ‘missing not at random’ (MNAR) and ‘missing at random’ (MAR) growth model comparison with a buprenorphine/naloxone clinical trial. Addiction. 2015;110(1):51–8. http://doi.org/10.1111/add.12714

22 Rubin DB. Multiple imputation after 18+ years. J Am Stat Assoc 1996; 91(434): 473–489. DOI: 10.2307/2291635

23 Steptoe A, Breeze E, Banks J, Nazroo J. Cohort profile: the English longitudinal study of ageing. Int J Epidemiol. 2013;42(6):1640–8. http://doi.org/10.1093/ije/dys168

24 Klaus D, Engstler H, Mahne K, Wolff JK, Simonson J, Wurm S, et al. Cohort Profile: The German Ageing Survey (DEAS). Int J Epidemiol. 2017;46(4):1105. http://doi.org/10.1093/ije/dyw326

25 Zhu Y, Pan D, He L, Rong X, Li H, Li Y, et al. China Registry Study on Cognitive Impairment in the Elderly: Protocol of a Prospective Cohort Study. Front Aging Neurosci. 2021;13:797704. http://doi.org/10.3389/fnagi.2021.797704

26 Gliklich R, Dreyer N, and Leavy M. Registries for evaluating patient outcomes: a user’s guide. Third edition (2014). Available at: http://www.effectivehealthcare.ahrq.gov/registries-guide-3.cfm.

27 Zhang Y. L. Multi center case registration of compound cistanche yizhi capsule in the treatment of vascular dementia based on the real world. (2021) Available at: http://www.chictr.org.cn/showproj.aspx?proj=139594.

28 Feng L, Kong L, Dong X, Lai X, Zhang D, Ren B, et al. China Stroke Registry for Patients With Traditional Chinese Medicine (CASES-TCM): Rationale and Design of a Prospective, Multicenter, Observational Study. Front Pharmacol. 2021;12:743883. http://doi.org/10.3389/fphar.2021.743883

29 Ong WY, Wu YJ, Farooqui T, Farooqui AA. Qi Fu Yin-a Ming Dynasty Prescription for the Treatment of Dementia. Mol Neurobiol. 2018;55(9):7389–400. http://doi.org/10.1007/s12035-018-0908-0

30 Jian-bo H, Guang-ji Z. Origin and innovative development of holism of traditional Chinese medicine. China Journal of Traditional Chinese Medicine and Pharmacy 2020; 35(01): 35–38. DOI: CNKI:SUN:BXYY.0.2020-01-006

31 Gaglio B, Henton M, Barbeau A, Evans E, Hickam D, Newhouse R, et al. Methodological standards for qualitative and mixed methods patient centered outcomes research. BMJ. 2020;371:m4435. http://doi.org/10.1136/bmj.m4435

32 Ruijin Q, Chen Z, Changming Z, et al. Application of mixed methods research in individualized therapeutic evaluation of clinical trials of traditional Chinese medicine. Chinese Journal of Evidence-Based Medicine 2020; 20(08): 973–978. DOI: 10.7507/1672-2531.201911079

33 Hongling C, Kaiwen N, Lin Z, et al. The Application of Mixed Methods Research in Real World Studies. Chinese Journal of Evidence-Based Medicine 2018; 18(11): 1203-1206. DOI:10.7507/1672-2531.201808035

34 Prins ND, Scheltens P. White matter hyperintensities, cognitive impairment and dementia: an update. Nat Rev Neurol. 2015;11(3):157–65. http://doi.org/10.1038/nrneurol.2015.10

35 Pasi M, van Uden IW, Tuladhar AM, de Leeuw FE, Pantoni L. White Matter Microstructural Damage on Diffusion Tensor Imaging in Cerebral Small Vessel Disease: Clinical Consequences. Stroke. 2016;47(6):1679–84. http://doi.org/10.1161/STROKEAHA.115.012065

36 Ashton NJ, Leuzy A, Lim YM, Troakes C, Hortobágyi T, Höglund K, et al. Increased plasma neurofilament light chain concentration correlates with severity of post-mortem neurofibrillary tangle pathology and neurodegeneration. Acta Neuropathol Commun. 2019;7(1):5. http://doi.org/10.1186/s40478-018-0649-3

37 Hoyer-Kimura C, Konhilas JP, Mansour HM, Polt R, Doyle KP, Billheimer D, et al. Neurofilament light: a possible prognostic biomarker for treatment of vascular contributions to cognitive impairment and dementia. J Neuroinflammation. 2021;18(1):236. http://doi.org/10.1186/s12974-021-02281-1

38 Walker KA, Gottesman RF, Wu A, Knopman DS, Gross AL, Mosley TJ, et al. Systemic inflammation during midlife and cognitive change over 20 years: The ARIC Study. Neurology. 2019;92(11):e1256–67. http://doi.org/10.1212/WNL.0000000000007094

39 Erhardt EB, Adair JC, Knoefel JE, Caprihan A, Prestopnik J, Thompson J, et al. Inflammatory Biomarkers Aid in Diagnosis of Dementia. Front Aging Neurosci. 2021;13:717344. http://doi.org/10.3389/fnagi.2021.717344

